# Age and sex impact resting-state EEG aperiodic activity beyond symptom associations in persistent post-concussive symptoms

**DOI:** 10.64898/2026.10.22.26366086

**Authors:** M. Louras, G. D’Archambeau, M. Meys, G. Martens, J-F. Kaux, J. Annen, A. Thibaut, N. Lejeune

## Abstract

1.

**Objective:** to investigate whether resting-state EEG aperiodic parameters (i.e., offset and exponent) are associated with symptom burden and demographic factors in patients with persistent post-concussive symptoms (PPCS) and differ from healthy controls.

**Methods:** In this cross-sectional observational study, resting-state eye-closed high-density EEG was recorded in patients with PPCS and in age-matched healthy controls. Aperiodic parameters were extracted from EEG power spectra. In patients, hierarchical multiple linear regression models included age and sex as a base model, with additional demographic and clinical symptoms entered separately. Patient-control differences were assessed using multiple linear regression adjusted for age and sex.

**Results:** Seventy patients with PPCS (54 female; 39.57±12.21 y.; 109.5 [76.5; 196] days post-concussion) recruited from a concussion care pathway and 38 healthy controls (17 female; 39.61±13.57 y.) were included. In patients, younger age and male sex were associated with higher offset and exponent values (all *p* ≤.014). None of the additional demographic and clinical variables improved model fit beyond age and sex (all *p*≥.143). After adjustment for age and sex, neither offset (*p*=.898) nor exponent (*p*=.100) differed between patients and controls.

**Conclusion:** Resting-state aperiodic EEG parameters were consistently associated with age and sex, but not with self-reported symptom burden or clinical characteristics in patients with PPCS. Not accounting for these factors may generate apparent group differences attributable, at least in part, to demographic variability.

**Significance:** Age and sex are important sources of inter-individual variability in resting-state aperiodic EEG activity and should be systematically considered when investigating such features.

**Highlights:**

- Age and sex significantly impact resting-state EEG aperiodic activity.
- Aperiodic EEG parameters were not associated with symptom burden in persistent post-concussive symptoms.
- Aperiodic offset and exponent did not differ between patients with persistent post-concussive symptoms and healthy controls after adjustment for age and sex.

## 1. Introduction

Concussion, or mild traumatic brain injury (mTBI), is a major public health issue, with global estimates reaching 56 million annual cases (Dewan et al., 2018). While most patients recover spontaneously within days or weeks, a substantial proportion develop persistent post-concussive symptoms (PPCS), with prevalence estimates ranging from 2% to 55%, and frequently reported around 30% (Cancelliere et al., 2023). PPCS commonly include somatic symptoms (e.g., headaches, sensitivity to noise/light), emotional symptoms (e.g., anxiety, depression), cognitive impairments (e.g., executive functioning, attention, working memory deficits) and sleep-related complaints (Mayer et al., 2017). PPCS may persist for months or even years following the injury and can significantly impair daily functioning, quality of life, and return to work (Gaudette et al., 2022).

Despite, the neurophysiological mechanisms underlying PPCS remain poorly understood, limiting the development of objective biomarkers and targeted interventions.

Resting-state electroencephalography (EEG) provides a non-invasive approach to investigate brain function and its relationship to behavior. Traditionally, EEG signals have been characterized in terms of periodic neural oscillations, divided in delta (1–4 Hz), theta (4–8 Hz), alpha (8–13 Hz), beta (13–30 Hz), and gamma (>30 Hz) frequency bands. These oscillations reflect coordinated neuronal activity across large-scale brain networks and have commonly been the primary focus of EEG research in neurological disorders (Formica et al., 2025). In the context of concussion, alterations in spectral power across frequency bands have been reported in the chronic phase post-mTBI, with patients exhibiting increased theta power, reduced alpha power, and decreased beta-band interhemispheric coherence compared to healthy controls (Lewine et al., 2019). However, the individual EEG measures showed limited ability to discriminate patients with mTBI from healthy controls, with classification accuracy below 60%, underscoring the limited specificity of these biomarkers.

Beyond canonical oscillatory activity, the EEG power spectrum also contains a broadband aperiodic component, often described as a 1/f-like signal, where power decreases with increasing frequency according to an approximate power law (Freeman et al., 2003). This aperiodic activity has been proposed to reflect fundamental aspects of neural population dynamics, including asynchronous synaptic activity and excitation-inhibition (E/I) balance (Gao et al., 2017; Miller et al., 2009). Acute concussion induces a neurometabolic cascade characterized by excessive glutamate release, altered ionic homeostasis, and alterations in excitatory and inhibitory neurotransmission (Giza & Hovda, 2014). These processes strongly suggest an alteration of cortical E/I balance during the acute phase. As concussion is known to disrupt excitatory and inhibitory neurotransmission during the acute phase, and because aperiodic EEG activity has been proposed as an indirect marker of cortical E/I balance, investigating aperiodic EEG parameters may provide novel insights into persistent neurophysiological alterations underlying PPCS.

The aperiodic component can be characterized by two parameters: the aperiodic offset, reflecting broadband shifts in spectral power, and the aperiodic exponent, describing the rate at which power decreases with increasing frequency and corresponding to the negative slope of the power spectrum in log-log space (Figure 1) (Donoghue et al., 2022). Donoghue and colleagues demonstrated that the aperiodic component accounts for a substantial proportion of the EEG spectrum and that changes previously attributed to oscillatory activity may, in part, reflect alterations in this broadband component (Donoghue et al., 2020). These findings highlight the importance of explicitly parameterizing the aperiodic signal when interpreting EEG spectra.

**Figure 1.**
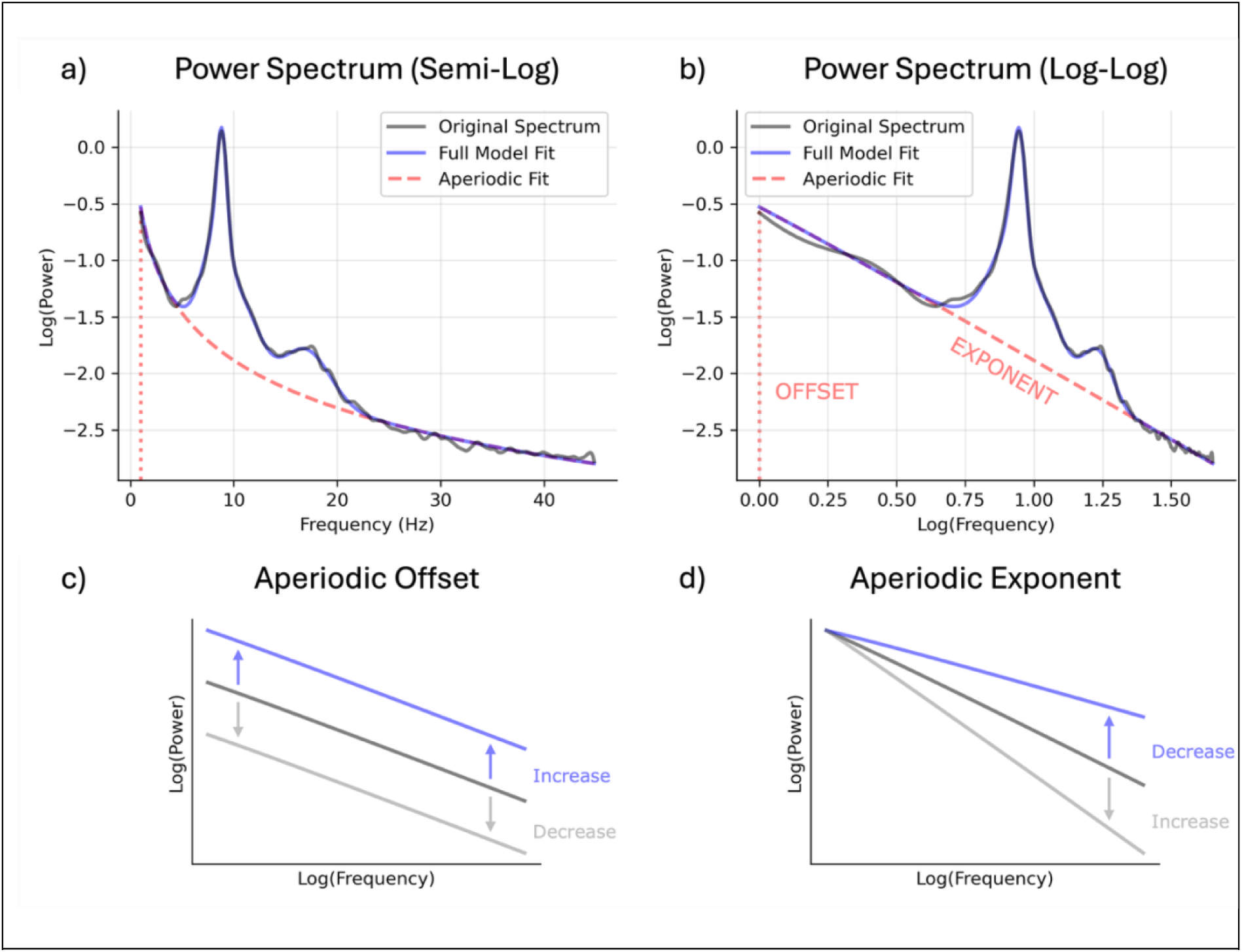
Illustration of the decomposition of the EEG power spectrum and the effects of the aperiodic parameters (offset and exponent) on the spectral profile. Panels a) and b) display the decomposition of the power spectrum of a representative participant. The empirical spectrum (gray line) is modeled as an aperiodic component (red dashed line) with superimposed oscillatory peaks, forming the full model fit (blue line). Panel a) presents the spectrum on the conventional semi-log scale with linear frequency axis, clearly showing oscillatory peaks in alpha and beta frequency ranges. Panel b) shows the same spectrum in log-log space, in which the aperiodic component becomes approximately linear and can be parameterized by an offset and exponent value, corresponding to the intercept and negative slope of the aperiodic fit, respectively. Panels c) and d) schematically illustrate how changes in the aperiodic parameters affect the power spectrum: changes in offset produce vertical shifts of the spectrum, whereas changes in exponent alter the steepness of the aperiodic slope.

Recent studies in spectral parameterization have shown increased interest in the aperiodic component as a potential biomarker of brain function. Alterations in aperiodic EEG activity have been reported across a number of neurological and psychiatric conditions, including Alzheimer’s disease (Wang et al., 2023), Parkinson’s disease (Monchy et al., 2024), schizophrenia (Peterson et al., 2023), ADHD (Robertson et al., 2019), major depressive disorder (Zandbagleh et al., 2024), disorders of consciousness (Maschke et al., 2025) and primary insomnia (Bai et al., 2025). Together, these findings suggest that aperiodic EEG parameters may capture fundamental alterations in cortical population dynamics that are shared across neurological and psychiatric disorders. However, their relevance to concussion remains largely unexplored. To our knowledge, evidence regarding aperiodic EEG alterations following concussion is currently very limited, with no established evidence that these parameters are sensitive to concussion-specific related outcome across different stages of recovery.

PPCS may reflect complex and heterogeneous alterations in brain function that are not captured by conventional clinical assessment or structural neuroimaging. Identifying objective neurophysiological measures associated with PPCS could therefore contribute to a better understanding of the neurobiological processes associated with the persistence of symptoms. Resting-state aperiodic EEG parameters represent promising candidate measures, as they provide non-invasive indices of brain activity that can be obtained from brief recordings and have been associated with alterations in brain physiology across neurological and psychiatric conditions. Investigating these parameters may therefore help determine whether they capture neurophysiological alterations associated with persistent post-concussive symptomatology.

Therefore, the primary analysis of the present study examined the relationship between resting-state EEG aperiodic activity and overall self-reported symptom burden in patients with PPCS. We hypothesized that greater symptom burden would be associated with higher aperiodic exponent and offset values, potentially reflecting altered cortical excitation-inhibition dynamics and broadband cortical activity. A secondary analysis compared aperiodic EEG parameters between patients with PPCS and healthy controls, with the hypothesis that patients would exhibit higher aperiodic exponent and offset values.

## 2. Material s methods

### 2.1 Study design

This single-center, cross-sectional observational study was designed to investigate the association between aperiodic EEG parameters (i.e., offset and exponent) and symptom severity in patients with PPCS, as well as differences in these parameters between patients with PPCS and healthy controls. Data collection was conducted at the University Hospital of Liège, Belgium, between May 2022 and February 2026.

### 2.2 Study participants

#### 2.2.1. Patients

Participants were recruited through a structured concussion care pathway managed by the University and University Hospital of Liège. Patients attending clinical appointments within this care pathway were systematically informed about the present study and invited to participate. The study used a convenience sampling approach, with the sample size determined by the number of eligible participants recruited through the clinical care pathway during the study period.

The diagnosis of concussion was made by a medical doctor according to the American Congress of Rehabilitation Medicine (ACRM) criteria (Silverberg et al., 2023). PPCS were defined by the persistence of symptoms beyond four weeks combined with a clinically significant symptom burden, as indicated by a Rivermead Post-Concussion Questionnaire (RPQ) score ≥ 16. Additional inclusion criteria comprised an age between 18 and 65 years and PPCS between four weeks and 12 months post-injury.

Exclusion criteria comprised a history of untreated neurological or psychiatric disorders within the six months preceding inclusion, evidence of an acute traumatic intracranial lesion on post-injury CT imaging, current untreated sleep disorders (e.g., sleep apnea), other severe medical conditions, self-reported alcohol or substance use disorders, previous participation in the concussion care pathway, and non French-speaking.

Potential participants received a detailed explanation of the study protocol by telephone prior to enrollment. Written informed consent was subsequently obtained either electronically or on the day of the study visit. The study protocol received approval from the Hospitalo-Faculty Ethics Committee of the University Hospital of Liège (ID: 2020/104). All participants provided written informed consent in accordance with the Declaration of Helsinki.

#### 2.2.2. Healthy controls

Healthy controls were recruited as part of a separate prospective normative study designed to characterize multimodal neurophysiological and neuroimaging measures across the adult lifespan (18 to 90 years). Participants underwent a comprehensive assessment including self-report questionnaires, neuropsychological and cognitive evaluations (including the Montreal Cognitive Assessment), EEG recordings, MRI acquisitions, PET imaging, and additional clinical and behavioral examinations. For the present study, healthy controls were selected based on an age range matching the patient cohort (i.e., excluding healthy participants above 65 years old) and an RPQ score below 16 (i.e., asymptomatic controls). Because the available control sample contained fewer female participants than the patient cohort, matching on sex would have substantially reduced the sample size and statistical power. Therefore, all eligible controls within the specified age range were retained, and age and sex were included as covariates in all regression analyses to account for demographic differences between groups. All healthy participants were screened for the absence of neurological and psychiatric conditions and provided written informed consent prior to enrollment in accordance with the Declaration of Helsinki. The protocol was approved by the Hospitalo-Faculty Ethics Committee of the University Hospital of Liège (ID: 2020/104).

### 2.3. Self-reported questionnaires

The following questionnaires were collected on the day of participation either on paper or via Alchemer, a web-based survey collection tool (https://www.alchemer.com/).

#### 2.3.1. Rivermead Post-Concussion Symptoms Questionnaire (RPQ)

The RPQ is a 16-item self-report instrument designed to assess the severity of post-concussive symptoms following mild traumatic brain injury (King et al., 1995). Items are rated on a 5-point scale from 0 (not experienced) to 4 (severe problem), yielding a total score ranging from 0 to 64, with higher scores indicating greater symptom severity. A score of 16 or higher is commonly considered clinically significant for PPCS.

#### 2.3.2. Epworth Sleepiness Scale (ESS)

The ESS is an 8-item self-administered questionnaire assessing subjective daytime sleepiness (Johns, 1991). Each item is rated on a 4-point scale from 0 to 3, with total scores ranging from 0 to 24. Higher scores indicate greater daytime sleepiness.

#### 2.3.3. Pittsburgh Sleep Quality Index (PSQI)

The PSQI comprises 19 self-rated items used to assess subjective sleep quality and disturbances over the preceding month (Buysse et al., 1989). The different items contribute to seven component scores, each ranging from 0 to 3, which are summed to yield a global score ranging from 0 to 21. Higher scores indicate poorer sleep quality and greater sleep disturbance.

#### 2.3.4. Beck Depression Inventory (BDI)

The BDI is a 21-item self-report measure assessing the severity of depressive symptoms over the preceding two weeks (Beck et al., 1961). Each item is scored from 0 to 3, yielding a total score ranging from 0 to 63. Higher scores indicate greater depressive symptom severity.

#### 2.3.5. Beck Anxiety Inventory (BAI)

The BAI is a 21-item self-administered questionnaire assessing the severity of anxiety symptoms over the past week (Beck et al., 1988). Items are rated on a 4-point scale from 0 to 3, yielding a total score ranging from 0 to 63. Higher scores indicate greater anxiety symptom severity.

### 2.4. Electroencephalography (EEG)

#### 2.4.1. Data collection

High-density resting-state EEG data were acquired using an actiCHamp Plus system and associated BrainVision Recorder software (Brain Products GmbH, Gilching, Germany). EEG recordings were conducted using 128-channel passive sponge-based electrode nets (128Ch Wet-Sponge R-Net; Brain Products GmbH, Gilching, Germany) soaked in a saline (potassium chloride) solution, with cap sizes ranging from 53 to 59 cm, selected based on each patient’s head circumference. Prior to data collection, electrode impedances were checked, and additional saline was applied as needed to ensure optimal signal quality (i.e., impedance kept below 50 kΩ). Recordings lasted approximately 12 minutes, during which patients sat in a comfortable chair and were instructed to remain awake, relaxed, and as still as possible. Patients were asked to keep their eyes closed throughout the session and to minimize movement and muscle activity. Recordings took place in a quiet room with minimal external distractions. Patients were continuously monitored by the experimenter, who intervened if signs of drowsiness or sleep were detected either behaviorally or from the real-time EEG activity. Due to the susceptibility of EEG to motion artifacts, the recording duration was occasionally extended to ensure that at least 10 minutes of artifact-free data would be retainable for further analysis. EEG signals were online referenced to the vertex (Cz) and sampled at 500 Hz.

#### 2.4.2. Preprocessing

EEG data preprocessing was performed in MATLAB (version R2022a; The MathWorks, Inc., Natick, MA, USA) using a slightly modified version of the MOHAWK toolbox (https://github.com/srivaschennu/MOHAWK; based on methods described by (Chennu et al., 2014, 2017)); a semi-automated EEG cleaning pipeline designed to facilitate artifact detection, component classification, and preprocessing reproducibility. Raw EEG signals, originally recorded from 127 channels, were downsampled to 250 Hz, band-pass filtered between 0.5 and 70 Hz and notch filtered at 50 Hz (2000th-order Hamming-windowed finite impulse response zero-phase filter), and segmented into 5-second epochs that were baseline-corrected. Data were visually inspected and a maximum of 10% of channels (up to 12) were removed if they exhibited excessive noise. Epochs containing prominent artifacts were discarded, provided that at least 120 (equivalent to 10 minutes) remained available for analysis. Independent component analysis (ICA) was applied to identify and remove signal components reflecting ocular, muscular, and cardiac artifacts. Bad channels were then reconstructed using spherical spline interpolation, and signals were re-referenced to the average reference.

#### 2.4.3. Aperiodic parameter estimation

Power spectral estimates were obtained as produced by the MOHAWK pipeline, which uses functionalities from the FieldTrip toolbox (Oostenveld et al., 2011). These spectra were computed for each channel using a multitaper fast Fourier transform with discrete prolate spheroidal sequence tapers and ±0.3 Hz spectral smoothing across the 0.5–45 Hz frequency range.

Aperiodic parameters were estimated using the FOOOF package (version 1.1.1, (Donoghue et al., 2020)) in Python (version 3.13.5). The FOOOF (Fitting Oscillations & One-Over-F) algorithm separates broadband (1/f-like) activity from superimposed oscillatory peaks by modeling the latter as Gaussians. The aperiodic component *P_AP_*(*f*) of the power spectrum *P*(*f*) at frequency *f* is modeled as:

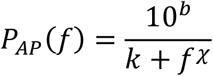

where *b* is the offset, *χ* is the exponent, and *k* is the knee parameter which captures the bend in the spectrum often present for broad frequency ranges (Miller et al., 2009). In case no bend is modeled (i.e., *k* = 0), and after log-transforming both sides of the equation, the aperiodic component takes the form of a linear function in log–log space, with *b* the intercept and *χ* the negative slope, as:

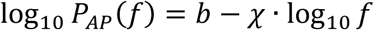

To accurately characterize the aperiodic component, it is important to appropriately model oscillatory peaks to ensure that they are properly removed from the spectra. Fitting too few or only very prominent peaks can leave genuine oscillatory activity in the aperiodic estimate, underfitting the model and inflating offset or exponent values, while fitting too many small peaks can absorb noise, overfitting the model and artificially flattening the aperiodic component.

To determine appropriate peak-modeling parameters for FOOOF, combinations for the maximum number of peaks (ranging from 3 to 6) and minimum peak height (0.05, 0.10, 0.20, and 0.30) were systematically evaluated. Resulting aperiodic parameter estimates remained largely stable across the considered range of peak-fitting settings, for which the model fit improvement reached a plateau with an increasing maximum number of peaks, which reduced when increasing the peak height threshold beyond a certain point. Based on this optimum, a maximum number of four peaks and a minimum peak height of 0.10 were selected, providing stable aperiodic estimates with high goodness-of-fit (mean R² = 0.98) and a physiologically plausible number of oscillatory peaks (mean = 3.74).

Because of the probable filter roll-off artifact near 0.5 Hz, which could bias the aperiodic parameter estimates, the frequency range for fitting was adjusted to 1–45 Hz. A fixed (no-knee) model was chosen given the relatively narrow frequency range of interest and simpler interpretation of the exponent parameter, alongside default values for the peak width limits (0.5–12 Hz) and relative peak detection threshold (2). The mean value for the aperiodic parameters was taken over all channels having a goodness-of-fit of R² ≥ 0.9.

### 2.5. Statistical analysis

#### 2.5.1. Normality and descriptive analyses

All statistical analyses were conducted using JASP (version 0.95.1). Continuous variables were assessed for normality using the Shapiro-Wilk test, complemented by visual inspection of Q-Q plots and histograms. Descriptive statistics are reported as mean ± standard deviation for normally distributed variables and as median with interquartile range for non-normally distributed variables.

Between-sex comparisons for demographic and clinical variables were performed using independent-samples *t*-tests when normality assumptions were met. For variables deviating from normality, the Mann-Whitney *U* test was applied. Categorical variables were compared using chi-squared (χ^2^) tests where appropriate. Statistical significance was set at α = .05 (two-tailed) for each analysis.

#### 2.5.2. Analyses in patients with PPCS

Associations between aperiodic EEG parameters and demographic and clinical characteristics were assessed in patients with PPCS using hierarchical multiple linear regression models. Separate models were fitted for offset and exponent as dependent variables.

For each model, age and sex were entered first as adjustment variables in the base model (M_0_). A variable of interest was then added in a second step (M_1_) to assess whether it explained additional variance in the aperiodic EEG parameter beyond age and sex. Variables of interest included years of education, time since injury (TSI), use of sleep medication, injury mechanism, and clinical measures of symptom burden (RPQ, ESS, PSQI, BDI, and BAI scores). Each variable was entered separately in the regression models. Injury mechanism was treated as a categorical variable, with road traffic accidents as the reference category and falls, sports-related injuries, work-related accidents, and assaults as the other categories.

The contribution of each variable was assessed by comparing the adjusted R^2^ of the base and full models and by examining the significance of the added predictor. This approach was used to determine whether demographic or clinical characteristics were associated with variability in aperiodic EEG parameters beyond the effects of age and sex.

#### 2.5.3. Patient - control comparisons

Group differences in aperiodic EEG parameters between patients with PPCS and healthy controls were assessed using separate multiple linear regression models, with group (PPCS vs. healthy controls) entered as the predictor of interest and age and sex included as adjustment variables. Separate models were fitted for offset and exponent, specified as follows:

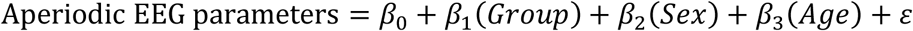

For all multiple linear regression models, model assumptions were assessed using Q-Q plots of standardized residuals and residuals-versus-predicted plots to evaluate the normality of residuals, linearity, and homoscedasticity. Multicollinearity was assessed using variance inflation factors (VIF). Missing questionnaire data occurred when participants did not complete the corresponding questionnaire. Analyses were conducted using complete cases for each regression model and no missing data were imputed. Multiplicity was addressed using Holm’s step-down procedure, applied separately within each model term across the two aperiodic components. Adjusted p-values are reported as p_Holm_.

## 3. Results

### 3.1. Participants

The patient screening and inclusion process is presented in Figure 2.

**Figure 2.**
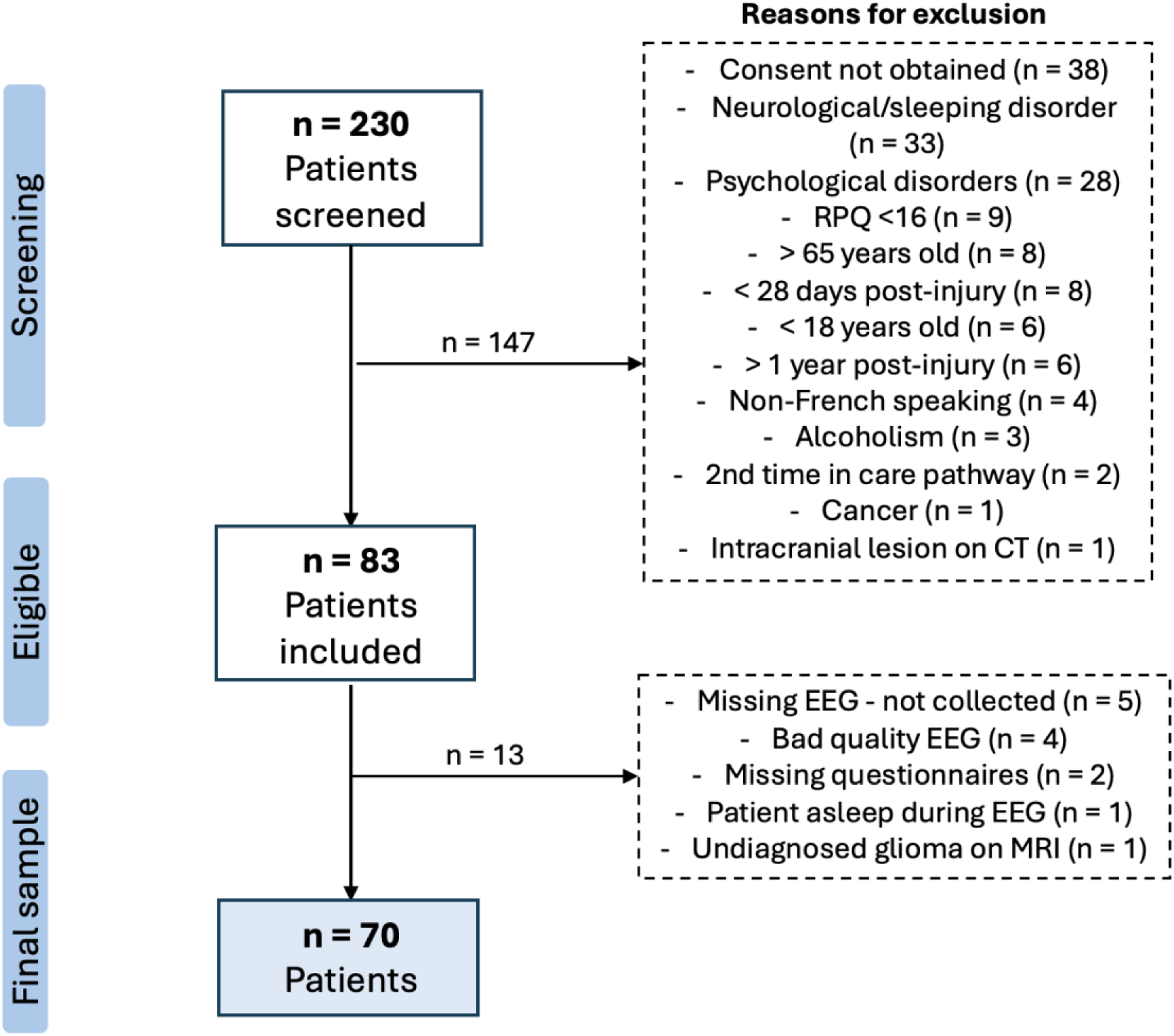
*Patient screening and inclusion flowchart.* Flow diagram illustrating the recruitment, eligibility assessment, and selection of patients with PPCS from initial screening to the final analytical sample. Reasons for exclusion at each stage are detailed in the corresponding boxes. Healthy controls are not represented in this flowchart, as they were recruited separately through an independent study, as described in the Methods section.

### 3.2. Patients and controls: demographical and clinical characteristics

The study included 70 patients with PPCS and 38 healthy controls. Demographic and clinical characteristics are summarized in Table 1.

**Table 1.**
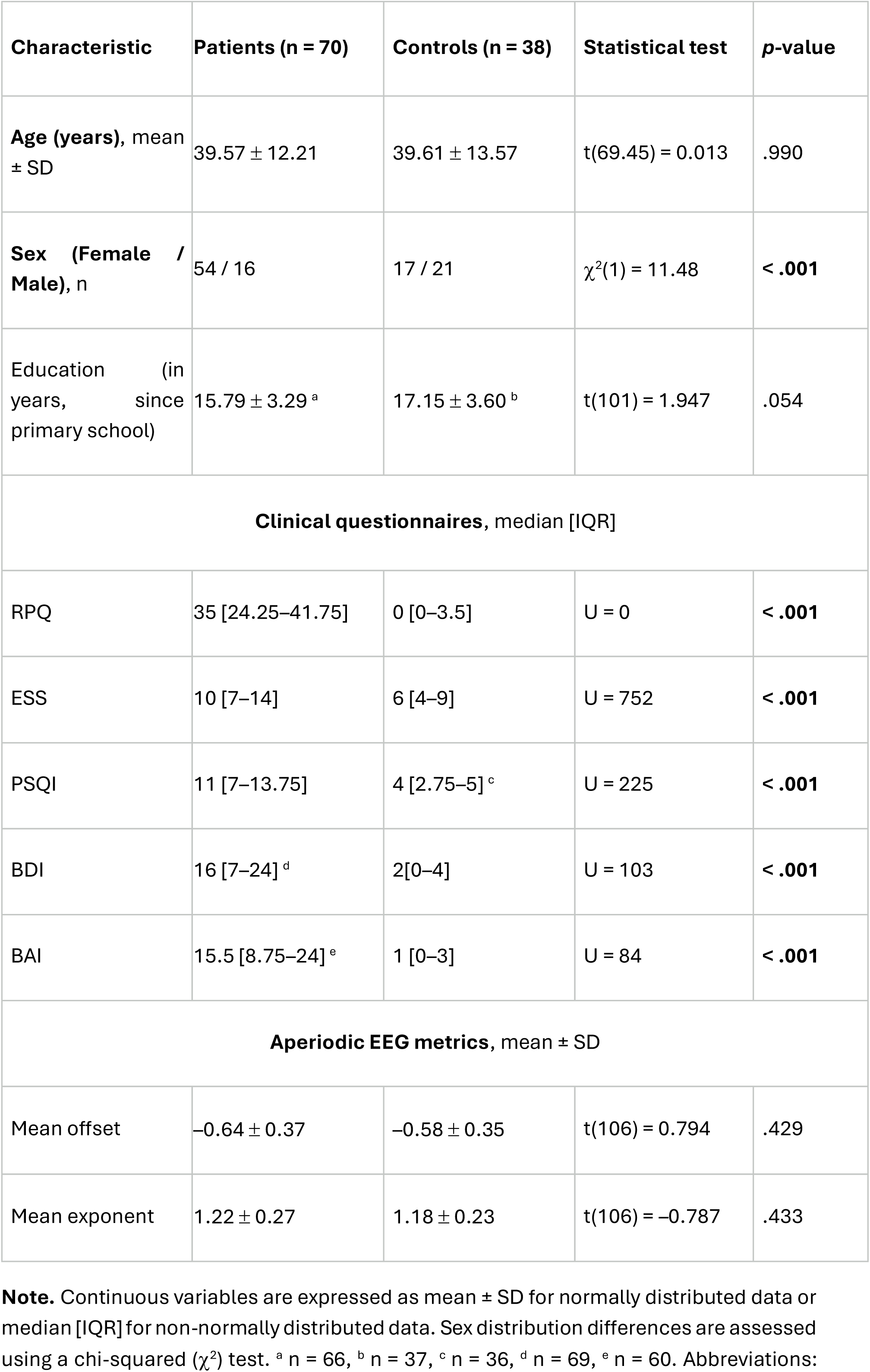
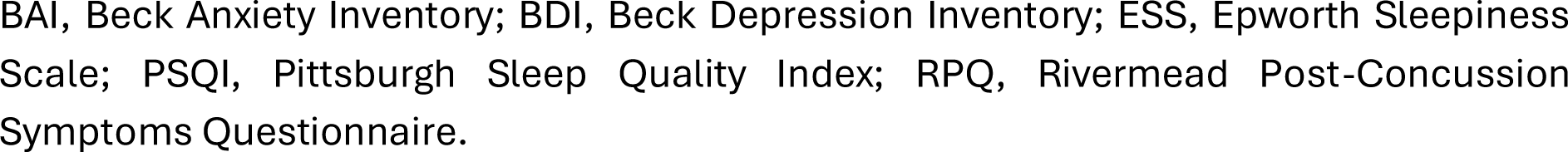
Demographic and clinical characteristics of patients with PPCS and healthy controls.

| Characteristic | Patients (n = 70) | Controls (n = 38) | Statistical test | p-value |
| --- | --- | --- | --- | --- |
| **Age (years)**, mean ± SD | 39.57 ± 12.21 | 39.61 ± 13.57 | t(69.45) = 0.013 | .990 |
| **Sex (Female / Male)**, n | 54 / 16 | 17 / 21 | $\chi^2(1) = 11.48$ | **< .001** |
| Education (in years, since primary school) | 15.79 ± 3.29 <sup>a</sup> | 17.15 ± 3.60 <sup>b</sup> | t(101) = 1.947 | .054 |
| **Clinical questionnaires**, median [IQR] |  |  |  |  |
| RPQ | 35 [24.25–41.75] | 0 [0–3.5] | U = 0 | **< .001** |
| ESS | 10 [7–14] | 6 [4–9] | U = 752 | **< .001** |
| PSQI | 11 [7–13.75] | 4 [2.75–5] <sup>c</sup> | U = 225 | **< .001** |
| BDI | 16 [7–24] <sup>d</sup> | 2[0–4] | U = 103 | **< .001** |
| BAI | 15.5 [8.75–24] <sup>e</sup> | 1 [0–3] | U = 84 | **< .001** |
| **Aperiodic EEG metrics**, mean ± SD | | | | |
| Mean offset | –0.64 ± 0.37 | –0.58 ± 0.35 | t(106) = 0.794 | .429 |
| Mean exponent | 1.22 ± 0.27 | 1.18 ± 0.23 | t(106) = –0.787 | .433 |

**Note.** Continuous variables are expressed as mean ± SD for normally distributed data or median [IQR] for non-normally distributed data. Sex distribution differences are assessed using a chi-squared ( $\chi^2$ ) test. <sup>a</sup> n = 66, <sup>b</sup> n = 37, <sup>c</sup> n = 36, <sup>d</sup> n = 69, <sup>e</sup> n = 60. Abbreviations:
BAI, Beck Anxiety Inventory; BDI, Beck Depression Inventory; ESS, Epworth Sleepiness Scale; PSQI, Pittsburgh Sleep Quality Index; RPQ, Rivermead Post-Concussion Symptoms Questionnaire.

Although patients and healthy controls were comparable in most socio-demographic factors, the patient group comprised a significantly higher proportion of females than the control group (77.1% vs. 44.7%, *p* < .001). Given this imbalance, additional analyses were conducted within the patient cohort to determine whether sex was associated with clinical or electrophysiological measures (Table 2).

**Table 2.**
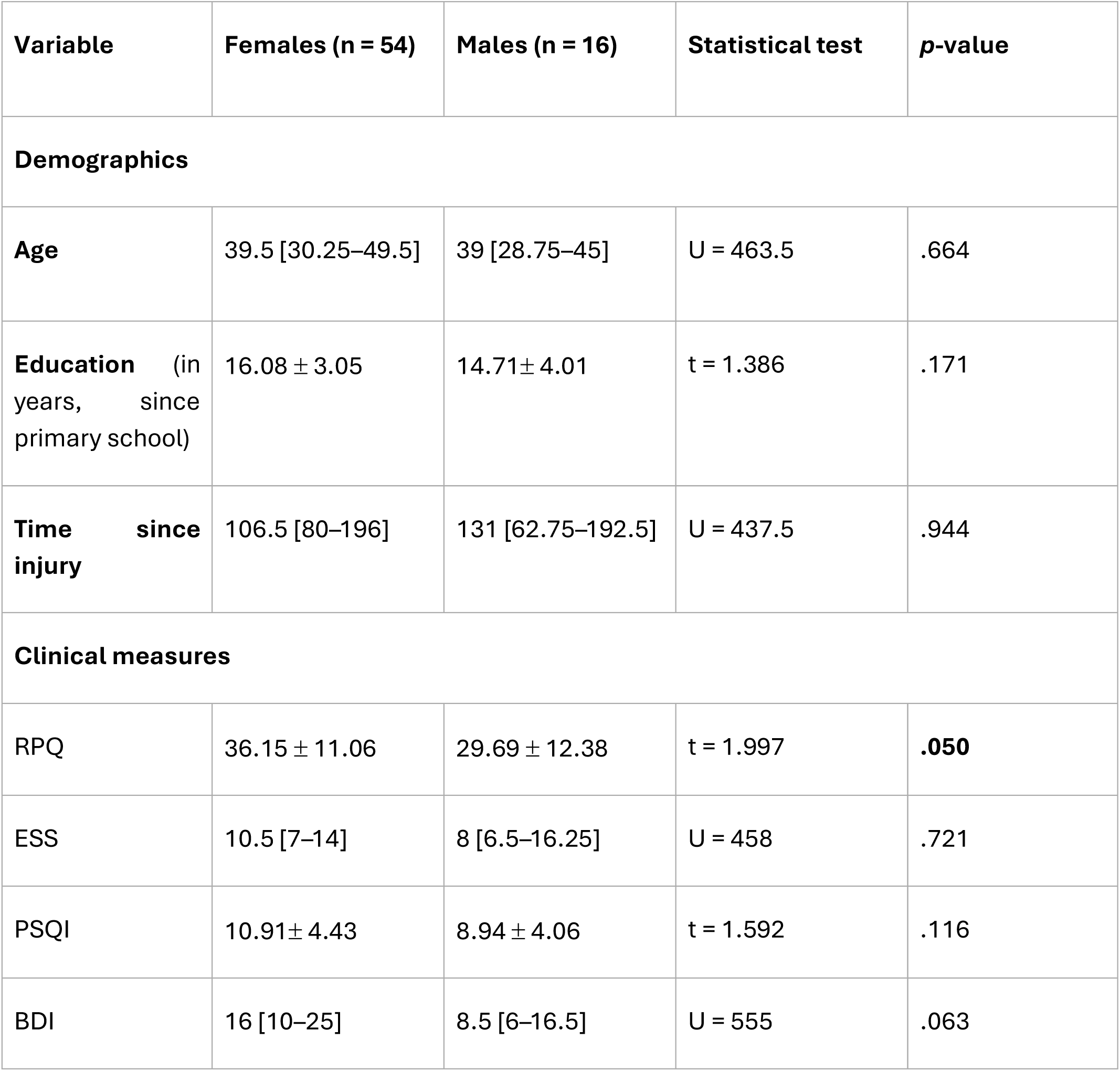
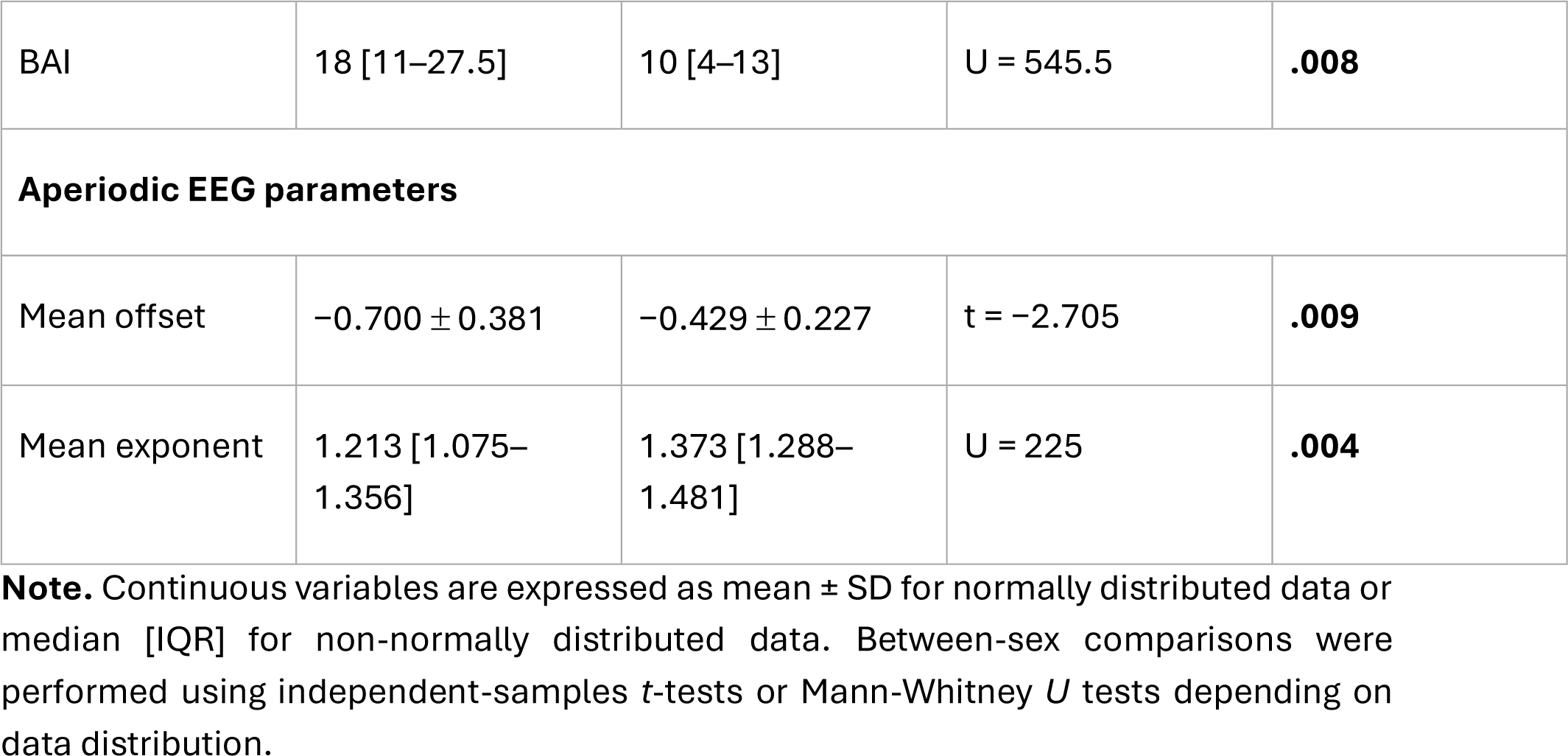
Sex differences in clinical symptom measures and aperiodic EEG parameters within the PPCS cohort.

| Variable | Females (n = 54) | Males (n = 16) | Statistical test | p-value |
| --- | --- | --- | --- | --- |
| **Demographics** |  |  |  |  |
| **Age** | 39.5 [30.25–49.5] | 39 [28.75–45] | U = 463.5 | .664 |
| **Education** (in years, since primary school) | 16.08 ± 3.05 | 14.71 ± 4.01 | t = 1.386 | .171 |
| **Time since injury** | 106.5 [80–196] | 131 [62.75–192.5] | U = 437.5 | .944 |
| **Clinical measures** |  |  |  |  |
| RPQ | 36.15 ± 11.06 | 29.69 ± 12.38 | t = 1.997 | **.050** |
| ESS | 10.5 [7–14] | 8 [6.5–16.25] | U = 458 | .721 |
| PSQI | 10.91 ± 4.43 | 8.94 ± 4.06 | t = 1.592 | .116 |
| BDI | 16 [10–25] | 8.5 [6–16.5] | U = 555 | .063 |
| BAI | 18 [11–27.5] | 10 [4–13] | U = 545.5 | **.008** |
| **Aperiodic EEG parameters** |  |  |  |  |
| Mean offset | $-0.700 \pm 0.381$ | $-0.429 \pm 0.227$ | t = -2.705 | **.009** |
| Mean exponent | 1.213 [1.075–1.356] | 1.373 [1.288–1.481] | U = 225 | **.004** |

**Note.** Continuous variables are expressed as mean ± SD for normally distributed data or median [IQR] for non-normally distributed data. Between-sex comparisons were performed using independent-samples *t*-tests or Mann-Whitney *U* tests depending on data distribution.

Within the PPCS cohort, no significant sex differences were observed for age, time since injury, daytime sleepiness (ESS), global sleep quality (PSQI) and depressive symptoms (BDI). However, significant sex-related differences emerged in some clinical and electrophysiological measures.

Regarding clinical measures, females reported higher post-concussive symptom severity on the RPQ (females: 36.15 ± 11.06; males: 29.69 ± 12.38, p =.050) and higher anxiety levels on the BAI (females: 18 [11–27.5]; males: 10 [4–13], p =.008).

Regarding aperiodic EEG parameters, significant demographic effects were observed. Male patients exhibited significantly higher mean offset values (females: −0.700 ± 0.381; males: −0.429 ± 0.227, p =.009) and higher mean exponent values, corresponding to steeper negative slopes (females: 1.21 [1.075–1.356]; males: 1.37 [1.288–1.481], p =.004) (Figure 3).

**Figure 3.**
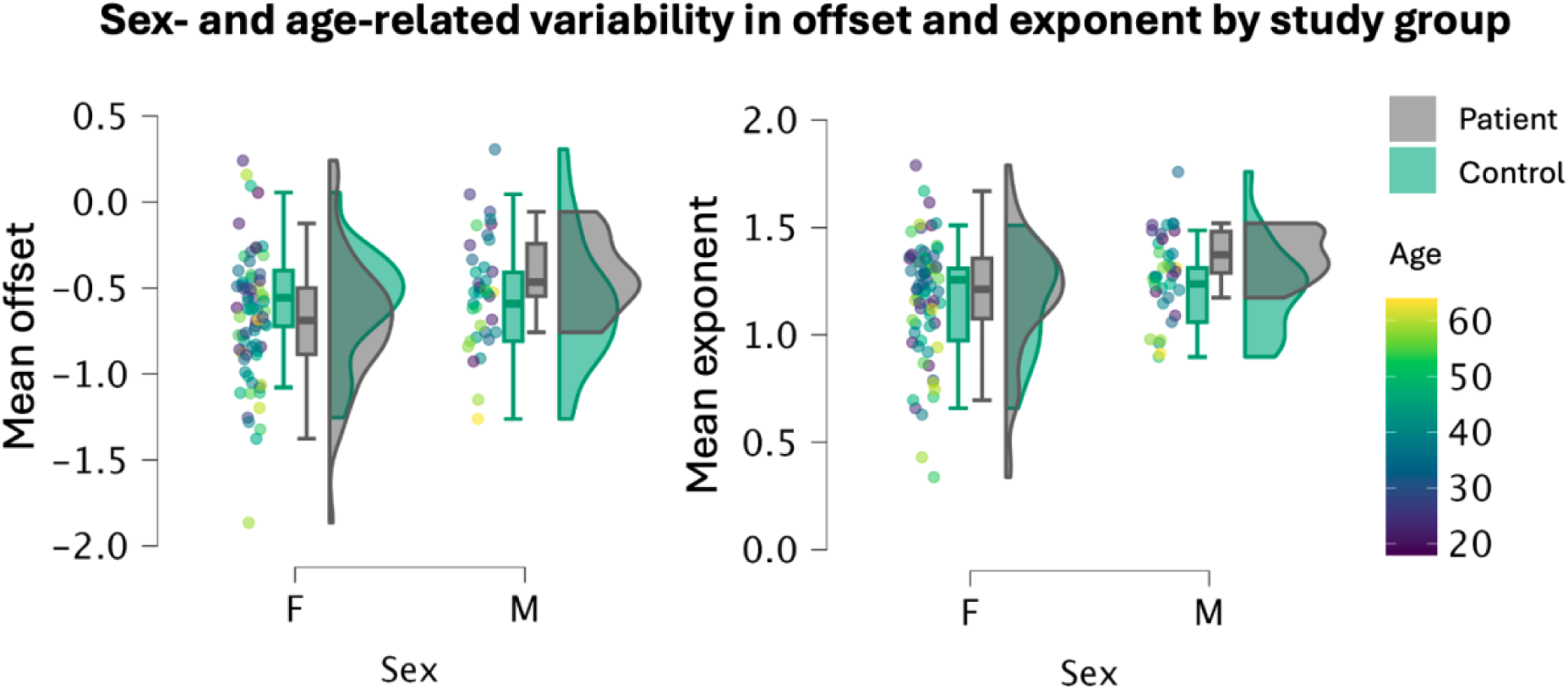
*Distribution of resting-state aperiodic EEG parameters according to sex, study group, and age.* Scatter plots, box plots, and violin plots of mean aperiodic offset (left) and exponent (right) for female (F) and male (M) participants in the PPCS (Patient) and control cohorts. Individual points represent single participants, with the color gradient reflecting age (18 to 65 years). Box plots depict the median, upper, and lower quartiles, while adjacent density plots show the overall data distribution.

In addition, age was significantly associated with both aperiodic parameters. Older age was associated with lower offset (β = −0.281, p =.014) and lower exponent values (β = −0.325, p =.004), indicating an age-related flattening of the aperiodic spectrum.

These findings further supported the inclusion of age and sex as adjustment variables in all subsequent analyses.

### 3.3. Primary outcome: aperiodic parameters in patients with PPCS

#### 3.3.1. Aperiodic offset

For the offset, the base models including age and sex explained between 13.0% and 15.2% of the variance (adjusted R^2^), depending on the available sample. Adding the additional variables resulted in adjusted values ranging from 11.9% to 15.3%. None of the additional predictors significantly improved the model beyond age and sex. Education showed the largest increase in adjusted R^2^ (from .137 to .153), although this improvement was not statistically significant (p =.143). Similarly, no significant additional contribution was observed for TSI, sleep medication use, injury mechanism, RPQ, BAI, BDI, ESS, or PSQI (Table 3).

**Table 3.** Hierarchical multiple regression analyses examining the contribution of demographic and clinical characteristics to aperiodic EEG parameters in patients.

| EEG parameter | Additional predictor | n | Adjusted R<sup>2</sup> – Base model | Adjusted R<sup>2</sup> – Full model | p-value of added predictor |
| --- | --- | --- | --- | --- | --- |
| Offset | Education | 66 | .137 | .153 | .143 |
|  | TSI | 70 | .152 | .150 | .359 |
|  | Sleep medication | 70 | .152 | .120 | >.430* |
|  | Injury mechanism | 70 | .152 | .144 | >.302* |
|  | RPQ | 70 | .152 | .144 | .528 |
|  | BAI | 60 | .130 | .119 | .616 |
|  | BDI | 69 | .151 | .138 | .874 |
|  | ESS | 70 | .152 | .148 | .402 |
|  | PSQI | 70 | .152 | .143 | .549 |
| Exponent | Education | 66 | .153 | .151 | .365 |
|  | TSI | 70 | .170 | .161 | .593 |
|  | Sleep medication | 70 | .170 | .147 | >.750* |
|  | Injury mechanism | 70 | .170 | .169 | >.427* |
|  | RPQ | 70 | .170 | .158 | .798 |
|  | BAI | 60 | .144 | .144 | .320 |
|  | BDI | 69 | .171 | .162 | .570 |
|  | ESS | 70 | .170 | .161 | .593 |
|  | PSQI | 70 | .170 | .158 | .796 |

**Note.** The base model ( $M_0$ ) included age and sex. Each additional predictor was entered separately in a second step ( $M_1$ ). Adjusted $R^2$ values are reported for the base and full models, and the $p$ -value corresponds to the additional predictor entered in $M_1$ . (\*) For categorical predictors, the reported $p$ -value corresponds to the omnibus test of the categorical variable.

#### 3.3.2. Aperiodic exponent

A similar pattern was observed for the exponent. The base models including age and sex explained between 14.4% and 17.1% of the variance, while the full models including the additional predictor explained between 14.4% and 16.9%. None of the additional variables significantly improved model fit beyond age and sex. Specifically, education, TSI, sleep medication use, injury mechanism, RPQ, BAI, BDI, ESS, and PSQI did not explain additional variance in exponent (Table 3).

Overall, these hierarchical regression analyses demonstrated that none of the demographic or clinical variables tested explained significant additional variance in aperiodic offset or exponent beyond age and sex. The consistency of the adjusted values across the base and full models further suggested that the variability in aperiodic EEG parameters was primarily accounted for by the demographic adjustment variables rather than by the additional clinical or injury-related characteristics examined.

### 3.4. Secondary outcome: patient-control comparison

Two multiple linear regression models were fitted to examine whether patient or control status was associated with offset and exponent after adjustment for age and sex (Table 4).

**Table 4.** Multiple linear regression models examining the association between patient– control group status and aperiodic offset or exponent.

| Outcome | Model F (*p*) | Adj. R<sup>2</sup> | Predictor | B (95% CI) | *p* | *p*<sub>Holm</sub> |
| --- | --- | --- | --- | --- | --- | --- |
| **Offset** | 4.475 (**.005**) | .089 | Group | −0.009<br>(−0.155, 0.137) | .898 | .898 |
|  |  |  | Age | −0.008<br>(−0.013, −0.002) | **.004** | **.008** |
|  |  |  | Sex | 0.150<br>(0.004, 0.297) | **.045** | **.045** |
| **Exponent** | 5.538 (**.001**) | .113 | Group | 0.085<br>(−0.017, 0.187) | .100 | .200 |
|  |  |  | Age | −0.005<br>(−0.009, −0.002) | **.004** | **.008** |
|  |  |  | Sex | 0.138<br>(0.036, 0.240) | **.009** | **.018** |

**Note.** Separate regression models were fitted for offset and exponent, with group (patients with PPCS vs. healthy controls), age, and sex entered simultaneously as predictors. Unstandardized regression coefficients (B), 95% confidence intervals (CI), and corresponding *p*-values are reported. Because offset and exponent were both evaluated as outcomes for each predictor, Holm's step-down procedure was applied within each predictor across the two outcomes (*m* = 2 per family).

Both regression models were statistically significant. However, after adjustment for age and sex, patient–control status was not associated with either offset (*p* = .898) or exponent (*p* = .100).

Consistent with the patient-only analyses, older age remained associated with lower offset and lower exponent values, corresponding to a flatter negative aperiodic slope, whereas male sex was associated with higher offset and higher exponent values, indicating a steeper negative aperiodic slope.

## 4. Discussion

In this study, we investigated the relationships between resting-state EEG aperiodic parameters (i.e., offset and exponent), and a multidimensional clinical presentation of PPCS. Across all investigated clinical domains, no significant associations were observed between clinical measures and EEG aperiodic parameters. Instead, age and sex consistently emerged as the strongest determinants of both aperiodic parameters. These demographic effects were stable across all regression models in patients and in the patient-control analyses.

Although the physiological interpretation of both parameters is still evolving (Porcaro & Bertoldo, 2026), the exponent has been proposed as a proxy of cortical excitation-inhibition balance (Gao et al., 2017), whereas the offset is thought to reflect overall broadband neural activity. Importantly, aperiodic parameters estimated using spectral parameterization approaches as implemented in FOOOF or its successor, specparam, appear to show generally good test-retest stability in healthy young adults, particularly the offset (McKeown et al., 2024), suggesting that they may capture relatively stable neurophysiological characteristics. Consistent with this interpretation, age and sex were the strongest predictors of the aperiodic component in our cohort, whereas self-reported symptom severity showed no independent associations after adjustment for these factors. Resting-state aperiodic EEG activity may therefore primarily reflect stable properties of cortical network organization rather than dynamic markers of current symptom burden.

### 4.1. Demographic factors as major determinants of aperiodic EEG activity

Among the investigated variables in patients with PPCS, age and sex appeared to be the most consistent correlates of resting-state aperiodic parameters. This pattern was maintained across all hierarchical regression models, irrespective of the additional demographic and clinical variables entered into the model.

The association with age was consistent with previous studies (Lago et al., 2026; Leroy et al., 2025; Merkin et al., 2023; Montemurro et al., 2024). In our sample, younger age was associated with both higher offset and higher exponent, corresponding to a steeper spectral slope. These findings align with the literature describing age-related reductions in exponent throughout adulthood, thought to reflect a shift in E/I balance toward inhibition, often accompanied by a decrease in offset, indicative of a reduction of overall cortical activity (Lago et al., 2026; Leroy et al., 2025; Merkin et al., 2023; Montemurro et al., 2024). However, age-related offset effects appear to be less consistent across studies, as Leroy et al. did not report such significant change in a large cohort of clinical EEG recordings (Leroy et al., 2025). Developmental studies further suggest that aperiodic parameters follow non-linear trajectories across childhood and adolescence, emphasizing that age-related influences likely reflect lifelong changes in cortical organization rather than disease-specific processes (Antúnez et al., 2025; Favaro et al., 2023; McSweeney et al., 2021).

Sex also showed a robust association with both aperiodic parameters. Men exhibited higher offset and exponent values than women, suggesting higher broadband neural activity and an E/I balance favoring inhibition. These associations remained significant across the models. Although evidence regarding sex effects remains less extensive than that for age, similar observations have been reported in developmental cohorts (Antúnez et al., 2025; McSweeney et al., 2021).

Taken together, the consistency of age- and sex-related effects across models highlights the importance of accounting for demographic characteristics when investigating aperiodic EEG parameters. Not accounting for these factors could potentially mask subtle disease-related effects or generate apparent group differences that are partly attributable to these demographical variables.

### 4.2. Clinical and injury-related factors beyond demographic characteristics

In addition, none of the additional demographic or clinical variables significantly improved our regression models. Specifically, education, time since injury, sleep medication use, injury mechanism, post-concussion symptom severity (RPQ), anxiety (BAI), depressive symptoms (BDI), daytime sleepiness (ESS), and sleep quality (PSQI) did not explain significant additional variance in either offset or exponent after adjustment for age and sex. The absence of associations between resting-state aperiodic parameters and these clinical outcomes is consistent with the multifactorial nature of PPCS, in which symptom severity emerges from complex interactions between biological, psychological and social factors (Iverson, 2019; Ponsford et al., 2019). The self-reported questionnaires included may capture broad clinical constructs that extend beyond resting-state cortical physiology, including emotional state, coping strategies, pain perception, and psychosocial influences. Future studies may benefit from combining aperiodic EEG analyses with more objective behavioral, cognitive, or physiological markers, such as psychomotor vigilance tasks, standardized neuropsychological assessments, actigraphy, or polysomnography. Longitudinal studies beginning in the acute phase could also help determine whether aperiodic parameters are more sensitive to early neurophysiological changes and examine how they evolve alongside symptoms over time.

### 4.3. Aperiodic parameters in patients and healthy controls

Neither offset nor exponent differed significantly between patients with PPCS and healthy controls after adjustment for age and sex, suggesting that the aperiodic EEG characteristics observed in the PPCS group were not clearly distinguishable from those of healthy individuals once demographic differences were accounted for. This is consistent with the only previous study investigating aperiodic EEG activity following mTBI, in which Nwakamma et al. likewise found no significant differences in offset or exponent between participants with mTBI and age- and sex-matched healthy controls within 14 days post-injury, in analyses additionally adjusted for age, sex, and years of education (Nwakamma et al., 2024). Together, this suggests that disease-related alterations in resting-state aperiodic EEG activity following concussion may be relatively subtle compared with the intrinsic biological variability associated with demographic factors. The lack of associations with self-reported clinical measures further suggests that aperiodic EEG parameters may capture aspects of resting-state cortical physiology that are not directly reflected by subjective symptom severity. This distinction may be particularly relevant when considering the potential of aperiodic parameters as objective neurophysiological markers in PPCS.

### 4.4. Broader methodological implications

Beyond PPCS, these findings also have broader methodological implications for the study of EEG aperiodic activity. Aperiodic parameters have been proposed as potential biomarkers across neurological and psychiatric conditions, yet demographic characteristics are not consistently handled across studies. Given that age and sex consistently emerged as the strongest determinants of offset and exponent in our cohort, demographic variability may contribute substantially to apparent disease-related differences, especially in conditions that are not equally distributed between sexes. Rather than questioning previous findings, our results emphasize the importance of carefully accounting for demographic factors during both study design (e.g., matching procedures) and statistical analyses (by accounting for these factors) to improve the sensitivity and reproducibility of future studies investigating aperiodic EEG biomarkers.

### 4.5. Limitations

This study has several limitations. First, most clinical variables relied on subjectively self-reported questionnaires, which do not constitute direct measures of neurophysiological state, sleep, or vigilance. Second, the sample size may have limited the statistical power needed to detect subtle neurophysiological alterations, while also precluding meaningful sex-stratified analyses as an alternative approach to address the imbalance between women and men. Third, recruitment through a specialized clinical pathway for PPCS may limit the generalizability of the results to the broader population of patients who sustained a concussion. Fourth, healthy controls were recruited as part of a separate normative study and were not prospectively matched to the PPCS cohort, which may have introduced unmeasured differences between groups despite adjustment for age and sex. Finally, the cross-sectional design and absence of pre-injury testing, including EEG recordings, preclude causal inference and do not allow determination of whether the observed aperiodic characteristics were present before injury.

Moreover, a methodological limitation of the current study concerns the inherent assumptions of spectral parameterization tools. Despite rigorous preprocessing and high signal quality, a small subset of participants (five individuals; 3 patients and 2 controls) exhibited suboptimal model fits, pragmatically defined as an R^2^ < .90 in more than 10% of the EEG channels (exceeding 12 channels). Visual inspection of the raw power spectrum densities confirmed that these suboptimal fits were not driven by artifacts but rather by an atypical physiological spectral architecture. These participants often presented a stable plateau extending from the alpha to the beta frequency bands, lacking the classic monotonic 1/f power decay over the frequency range of interest. This observation highlights a broader limitation of current spectral parameterization approaches. The assumptions underlying FOOOF may not adequately capture the full range of spectral characteristics encountered in healthy individuals or, particularly, in clinical populations, where spectral patterns may deviate substantially from those typically observed. Furthermore, the thresholds used to define acceptable model fits remain largely empirical and are not yet supported by clear consensus guidelines, introducing subjectivity into participant inclusion decisions.

Future research should therefore aim to better characterize atypical spectral profiles and determine whether they represent distinct neurophysiological phenotypes or limitations of existing parameterization models.

## 5. Conclusion

In patients with PPCS, resting-state EEG aperiodic parameters were more consistently associated with age and sex than with subjective clinical measures. No significant associations were observed with post-concussion symptom severity, subjective daytime sleepiness, global sleep quality, anxiety, or depression. Furthermore, aperiodic parameters did not significantly differ between patients with PPCS and healthy controls after adjustment for age and sex. Taken together, these findings suggest that resting-state EEG aperiodic parameters may not directly reflect subjective symptom burden in PPCS and highlight the importance of systematically accounting for demographic variables in studies investigating the EEG aperiodic spectral component.

## Data Availability

All data produced in the present study are available upon reasonable request to the authors

## Funding sources

The study was supported by the University and University Hospital of Liège, the Belgian National Funds for Scientific Research (FRS-FNRS) and the Leon Frédéricq Foundation.

M.L. is a Research Fellow of the F.R.S.-FNRS, G.M. is a Postdoctoral Researcher of the F.R.S.-FNRS, A.T. is a Research Associate of the F.R.S.-FNRS, and N.L. is a Clinical Researcher of the F.R.S.-FNRS. We are also highly grateful to the members of the NeuroRecovery Lab for their assistance in clinical evaluations, especially Nathalie Maquet. We are also thankful to the patients for agreeing to take part to this study.

## Contributions

M.L. and G.D. contributed equally to the conceptualization and methodological development of the study, performed the statistical analyses, and drafted the manuscript. M.M. contributed to the methodological development and analysis of the aperiodic EEG parameters, under J.A. supervision, and to manuscript revision. G.M., J.A., and J.-F.K. contributed to critical review and revision of the manuscript. A.T. and N.L. contributed to study conceptualization, supervision, and critical revision of the manuscript. All authors approved the final version of the manuscript and agreed to its submission.

## Declaration of AI

The authors used ChatGPT (OpenAI) to assist with language refinement and improve the clarity of selected sections. The authors then reviewed and edited the content and take full responsibility for the content of the publication.

## Data availability

The data supporting the findings of this study are not publicly available because they contain sensitive clinical and neurophysiological information and are subject to institutional and data protection restrictions. Data may be available from the corresponding author upon request and subject to appropriate ethical and data-sharing approvals.

